# A public health approach to cervical cancer screening in Africa through community-based self-administered HPV testing and mobile treatment provision

**DOI:** 10.1101/2019.12.19.19015446

**Authors:** Miriam Nakalembe, Philippa Makanga, Andrew Kambugu, Miriam Laker-Oketta, Megan Huchko, Jeffrey Martin

## Abstract

**Background:** Sub-Saharan Africa bears the world’s highest incidence of cervical cancer. To address the lack of widespread screening and treatment that contributes to this burden, the World Health Organization (WHO) recommends that low-resource countries adopt simplified protocols for screening directly coupled with treatment. The WHO recommendations present an opportunity — akin to what has been done for HIV care in Africa — for a true public health approach to cervical cancer control in resource-poor settings. We evaluated the feasibility of such a public health approach to cervical cancer that features community-based self-administered HPV screening and mobile treatment provision.

**Methods:** In two rural districts of western Uganda, we first trained Village Health Team members (VHTs, also known as Community Health Workers) in a one-day session in the fundamental aspects of cervical cancer and its prevention. We then provided guidance to the VHTs to mobilize adult women from different communities within the district to attend a one-day HPV screening fair at a central location in their respective community. On the day of the fair, the study team and VHTs provided educational talks and instructions for self-collection of a vaginal sample. The samples were subsequently tested for high risk HPV (hrHPV) E6/E7 mRNA using the APTIMA^®^ platform. Women who tested positive for hrHPV were re-contacted and referred for treatment with cryotherapy at a mobile treatment unit in their community. Visual assessment with acetic acid was used to guide suitability for cryotherapy in the mobile treatment unit versus further referral to a larger facility for a loop electrosurgical excision procedure (LEEP).

**Results:** Between March and November 2016, 2,142 women attended a health fair in one of 24 communities in rural Western Uganda and expressed interest in being screened for cervical cancer; 1902 were eligible for cervical cancer screening of which 1,892 (99.5%) provided a self-collected vaginal sample. The median age of those screened was 34 years (IQR: 28-40), HIV prevalence was 11%, and most (95%) had not been previously screened. Almost all women stated that they would perform the self-collection again and recommend it to a friend. Prevalence of any hrHPV mRNA was 21% (HPV-16, 6%; HPV-18/45, 1.9%). Among the 393 women with detectable hrHPV mRNA, 89% had their results transmitted to them, of whom 86% returned to the mobile treatment unit. At the mobile treatment, 85% of women underwent ablative therapy, with the remainder deferred either because of pregnancy (9.0%), need for LEEP (2.6%) or other reasons (3.3%).

**Conclusion:** A public health approach to cervical cancer screening, featuring community-based self-administered HPV testing and mobile treatment, was feasible and readily accepted by community women. The process is termed a “public health approach” because — as is the case for HIV care in the region — it explicitly concedes perfection at the individual level in deference to reaching a larger fraction of the population. The findings support further optimization and evaluation of this approach as a means of scaling up cervical cancer control in low resource settings. If resources for cancer control remain limited in sub-Saharan Africa, this public approach may offer one of the most efficient solutions for stemming the incessant tide of cervical cancer in the region.

## Introduction

Resource-limited regions bear the brunt of the burden of cervical cancer incidence and mortality worldwide. In 2018, there were an estimated 528,000 incident cases and 266,000 deaths caused by cervical cancer worldwide; with 85% and 90% of these in developing countries [1]. The situation is perhaps most dire in East Africa where age-standardized rates reached 40 new cases of cervical cancer diagnosed and 30 deaths from the disease per 100,000 person-years in women in 2018 [1] – this, by any definition qualifies as a public health problem. However, by contrast, incidence and mortality of cervical cancer in the U.S. 6.4/100,000 person-years and 1.9/100,000 person-years [1]. Most of this disparity is due to a lack of organized screening programs in Africa and hence lack of prevention of precancerous lesions becoming cancer [2, 3]. For example, in Uganda screening is erratic, opportunistic, and in some places absent translating into staggeringly low screening uptake, with rates as low as 4.8% in rural Uganda [4]. While human papillomavirus (HPV) vaccination has received a significant amount of public health attention and funding as the most cost-effective means to cervical cancer control, slow roll-out and a focus solely on adolescent girls in low-resource settings mean that screening programs will remain central to cervical cancer prevention in these populations for the foreseeable future.

The identification of HPV infection as a causal determinant of cervical cancer and subsequent development of sensitive and specific HPV diagnostic tests have been critical technologic advances that hold promise for decreasing worldwide cervical cancer burden. One of the most impactful and innovative applications of the HPV paradigm in cervical cancer is the development of tests that have been shown to have adequate sensitivity in vaginal specimens – with evidence that women can self-administer the collection of samples [5]. In places where these tests have been investigated, such as Uganda [6, 7] and Kenya [8], women prefer self-administered approach to provider-administered tests [9]. Offering self-administered tests in the community, without women having to travel to designated health facilities for them, should, in theory, increase their use and there is early evidence for this [10, 11]. Currently, the World Health Organization recommends the use of these tests with treatment based on a positive result without further confirmation [12]. However, there still remain challenges in getting treatment to those with positive tests [13, 14]. For example, requiring women tested in the community to travel great distances to facilities for treatment not surprising often results in substantial losses to follow-up and failure to administer therapy [11].

To address the public health problem of cervical cancer in sub-Saharan Africa, we sought to take advantage of the recent technologic advancements in HPV testing and coupled them with practical insights (e.g., mobile treatment provision) to develop a community-based multi-component “public health approach” to screening. We term this a public health approach because it brings services directly to the community, uses low cost processes at each step, and concedes perfection at the individual level in order to reach the greatest number of women. The approach is akin to the public health approach what was done for HIV care and prevention [15] – arguably one of the greatest biomedical successes of our generation. We set forth to determine the feasibility of such a public health approach in a demonstration project in Uganda.

## Materials and Methods

### Overall design

We evaluated the feasibility of a cervical cancer screening and treatment intervention consisting of Village Health Team (VHTM)-delivered community mobilization, self-collection of vaginal samples for HPV testing, and community-based mobile treatment for HPV-infected women in two rural districts of Uganda.

### Development of community-based message regarding cervical cancer screening

In each district, we sought permission from district leadership to carry out cervical cancer screening and employ VHTMs. With facilitation from the district leaders, we identified and invited key stakeholders that included the local council (LC) leaders, and community elders, to participate in the development of community-based campaign to encourage women to attend a fair and teach the women about the importance of cervical cancer screening. This process was led by Jive Media Africa, a health communication from based in South Africa, who used the Information-Motivation-Behavioral (IMB) model (Fisher and Fisher 1992) to develop posters, brochures, short oral narratives for the community mobilization (Figure 1).

**Figure 1.**
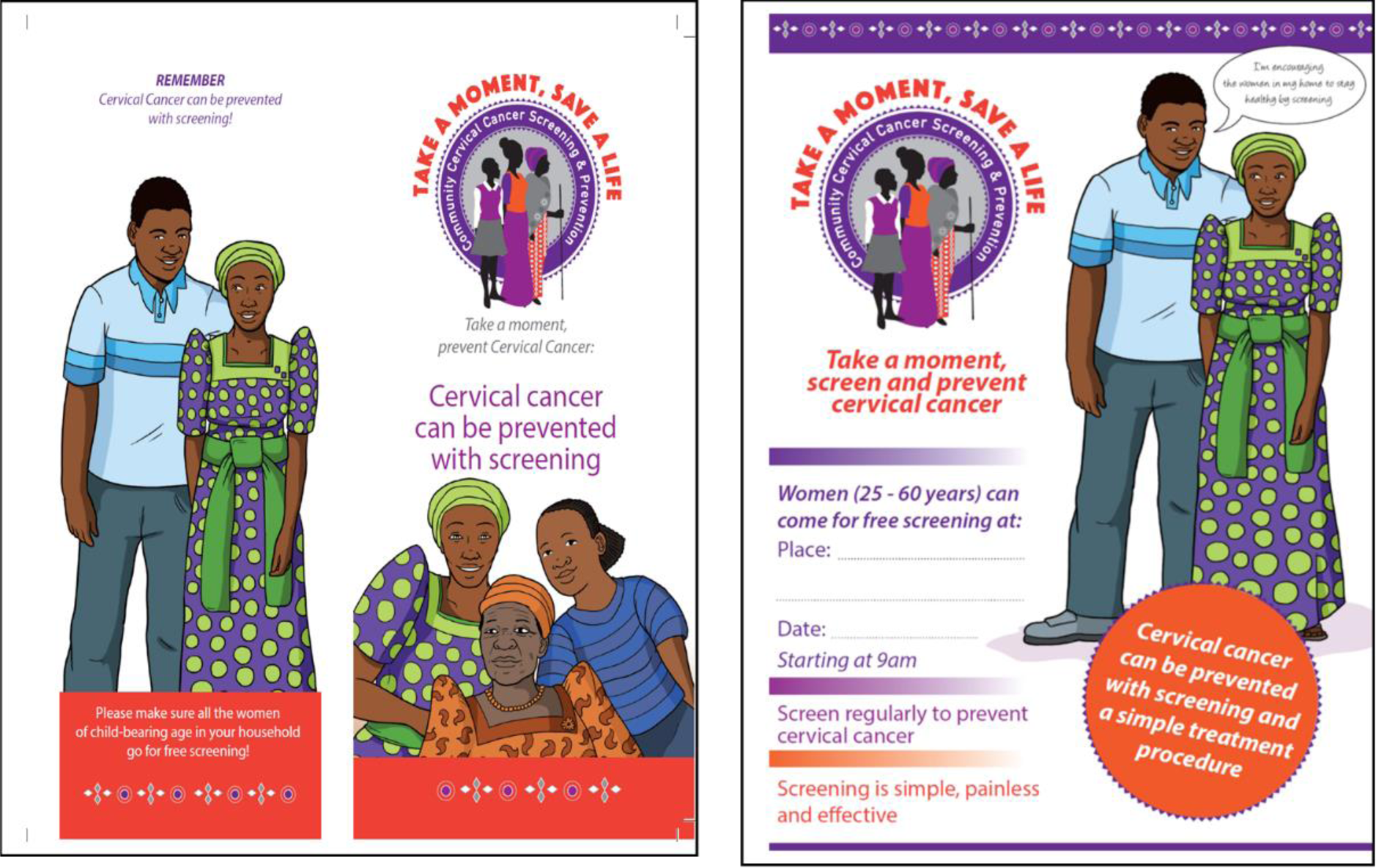
showing the posters that were used to invite women for screening indicating the date of the due date for the activity in the community campaign

### Community mobilization and study population

#### Target population and sampling

We sought to enroll women 25-60 years residing either in Kiboga or Kyankwanzi, two rural districts with mainly agricultural economies in western Uganda [16]. Within these districts, we randomly selected three sub-counties: Lwamata in Kiboga and Butemba and Ntwetwe in Kyankwanzi. In each sub-county, we purposively choose 5 to 9 parishes, which are administrative units with about 5-20 villages and a population of about 250-1000 people (we will refer to these as communities).

#### Community mobilization campaigns

Village Health Team Members (VHTMs) also known as community health extension workers (CHWs) are community members who receive training to provide basic education and care for selected health conditions. In Uganda, VHTs are trained in various areas to provide education, linkage to care and, in some cases, first-line treatment at the community level [17, 18]. The research staff hosted VHTMs for a one-day training session in their communities on the basics of cervical cancer, HPV self-testing and how to mobilize the community to attend a health fair dedicated to cervical cancer screening. We conducted a two-week mobilization campaign in each community, in which VHTMs informed residents about a health fair scheduled to take place within their communities on cervical cancer screening among women aged 25-60 years. The campaign did not mention that the fairs were being performed under the auspices of a research study. Occasionally, the VHTs went door-to-door to augment messaging provided through brochures, posters and oral narratives in market places, churches and trading centers.

### Community health fairs

The venues for the health fairs varied by community and included community centers or fields within the communities, around health centers or on school compounds. The VHTs selected the venues based on their centrality within the community and availability of space for the study activities. Upon arrival at each daily health fair, the study team, working alongside the VHTs, registered the women. At registration, the women provided their name, village of residence, age, parity and a brief gynecological history including presence of an abnormal vaginal discharge or bleeding and name. The study staff gave health talks to women in groups of 20-30 as they gathered at the health fairs. The health talks covered the burden of cervical cancer, risk factors, signs and symptoms, and prevention by screening as well as available treatment modalities for the pre-cancer stages. The sessions were interactive and facilitated with numerous visual aids (Figure 2). The women were provided with brochures containing information that had been discussed for their further reference. We conducted health fairs in each community location (represented by 5-20 villages within each parish) until a drop in participant numbers before moving on to the next community.

**Figure 2.**
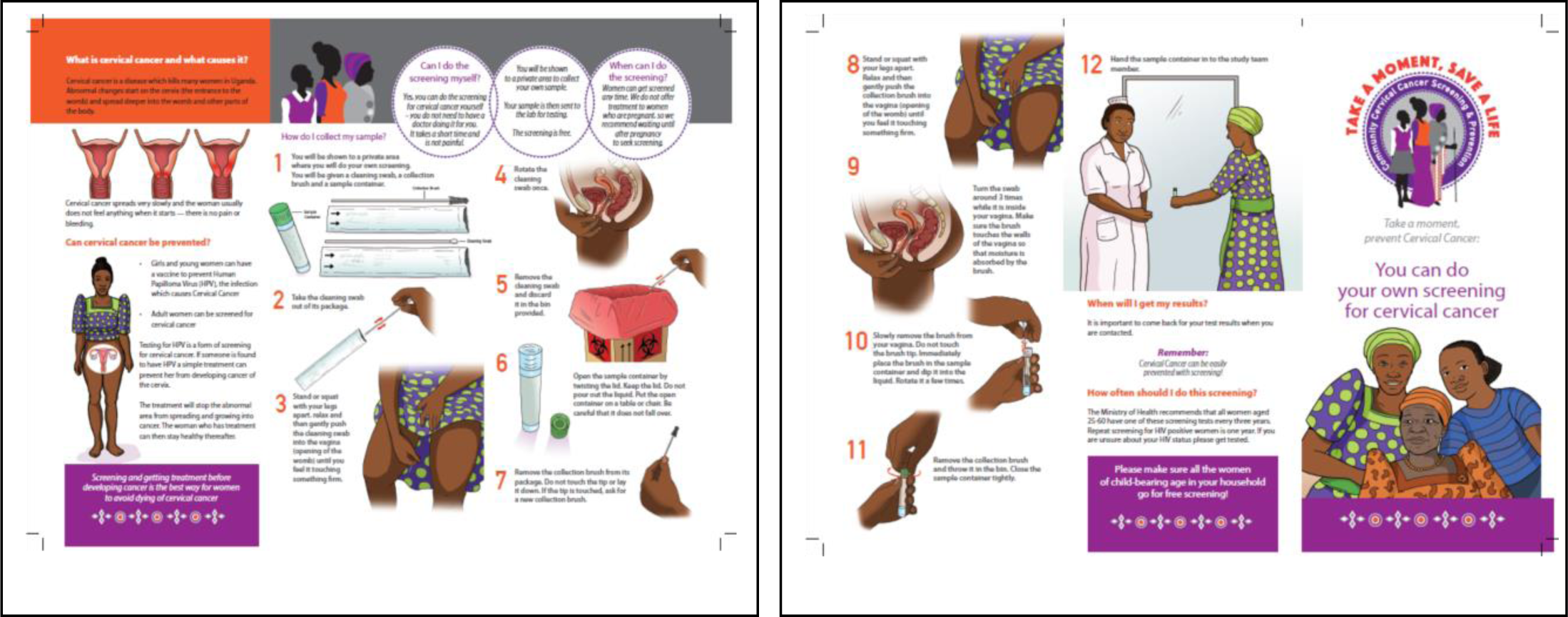
showing the Visual aids that were used at the screening venue.

### Study population

After the educational talk, all women age 25 to 60 years, were asked to provide written informed consent if they wished to participate. Women with a history of cervical cancer, symptoms and signs suggestive of cervical cancer (foul pus like vaginal discharge or bleeding with suspicious lesion on speculum exam), or hysterectomy were excluded. The study was approved by institutional review boards in both Uganda and the U.S., and all women provided written informed consent to participate.

## Measurements

### Vaginal specimen collection for HPV testing

The study staff together with the VHT carried out a demonstration on how to self-collect a vaginal sample for the HPV test using posters and brochures. They were also provided with a brochure specifically containing information and illustrations of how to self-collect the vaginal sample for their reference. Women were provided with an unopened collection kit (Aptima®, Hologic/Genprobe, Inc.) containing a cytobrush and a capped vial containing a collection medium. We provided floor-length screens inside tents for privacy in which women self-collected the vaginal specimen. Briefly, participants were instructed to place the brush inside the vagina until it met resistance and rotate it 3 times. The participants then placed the used brush into a collection vial with PreservCyt solution, broke the pre-scored handle, and closed the vial prior to returning it to the health worker. From the field, the specimens were transported at ambient temperature to the Translational Laboratory at the Infectious Diseases Institute, Makerere University. We have previously described the virological characterization of these women [19].

### Socioeconomic, demographic and clinical characteristics and self-collection experience

Prior to collection of the vaginal specimen, we used an interviewer-administered questionnaire to collect demographic, clinical, and reproductive health data as well as information on accessibility of the venue. After vaginal specimen collection, women were asked to assess the experience of their participation, instructions on self-testing, discomfort with the swab, whether they would test again as well as if they would recommend it to someone else. Originally, the responses were yes/no but when we observed scant variability we decided to change to a 5-point scale, which is what we will refer to in this current report. Information on preferred route of notification of results (short message service (SMS), phone call, home visit, or collection of results from their nearest health facility.), locator details that included phone numbers, physical address as well as contact VHT was collected.

### Result notification

Women were notified of their HPV results based on their preferred mode of notification. SMS notification was considered successful if transmission of text message was confirmed (i.e. phone was on, SIM card valid, line active). Phone and home visits done by the study staff were successful if participants were given their results directly by study staff. Participants were attempted to be contacted up to three times in a day over at least three different days within a period of 1-2 weeks. Clinic notification was considered successful if the participant returned to her nearest clinic to pick up her results by clinic staff. The staff put in more effort to find the hrHPV positive participants who were not reached based on their primary mode of notification. In case the primary mode was SMS or return to clinic, phone calls were also made. In case all this failed, the VHTMs physically tracked them and gave them appointments to come to the clinic and receive their results. They were finally declared lost if the VHT confirmed that they could not find them as happens in cases of relocation.

### Mobile treatment

Treatment for the HPV-infected women was offered using a mobile treatment strategy within the respective communities. We selected sites to provide treatment to optimize convenience for the women and, ultimately, attendance. We typically chose existing Health Center III or II facilities. At each site, the team provided a two to three-day window for participants to come for, anytime between 8am and 3pm. The VHTs participated with the team on that day to particularly search for the women who had appointment for that day but had not come. We term this component “mobile” treatment because the treatment team carried all their supplies (e.g., speculums, biopsies, vinegar as well as the cryotherapy treatment system). The treatment team consisted of a study doctor and study nurses as well as VHTs who assisted in the non-clinical organisation. Prior to treatment, the study staff collected information on pregnancy state as well as the last menstrual period, with urine pregnancy testing when indicated clinically or by history. (MOTI test^®^, Atlas Link, Beijing, China). Women who were found in menses were given another appointment for treatment at that facility within about 4 weeks while those found pregnant were told to get in touch with their VHT or study team 6 weeks after delivery. Visual assessment for treatment (VAT) was done to determine appropriate treatment. Cryotherapy was used for those women with lesions covering less than 75% of the cervix, no extension to the endocervix or vagina and no evidence of cancer. Although not required for real-world implementation, we also, as part of the research study, performed a biopsy on all women prior to cryotherapy. Women with lesions not amenable to treatment with cryotherapy (too large, entering the endo-cervical canal) were referred for at Mulago Hospital in Kampala (131 Kilometers). Women with lesions suspicious for cancer at the time of their VAT underwent biopsy. The women who were determined to have invasive cancer on biopsy were referred to the Uganda Cancer Institute. At the time of their appointment, all the women were informed that they would be facilitated with some transport refund including those who were referred to the treatment at the Uganda Cancer Institute.

### Statistical analysis

After determining the descriptive parameters for the population, we described the successful completion of each aspect of the cascade with percentages and 95% confidence intervals. We evaluated various participant sociodemographic and clinical characteristics for their association with successful notification of HPV test results and, among those who were HPV-infected, presentation for treatment. We used risk ratios as the measure of association and used log-binomial regression to estimate these adjusted for other factors. We used a directed acyclic graph (DAG) to depict background knowledge and inform variable selection in the multivariable regression models [20, 21] (Figure 3). Regarding compliance with the program’s directive among the HPV-infected women to attend a mobile treatment unit, near uniform attendance left us with insufficient variability to merit evaluation of determinants. All calculations were performed using Stata version 14.0 (Stata Corp., College Station, Texas).

**Figure 3.**
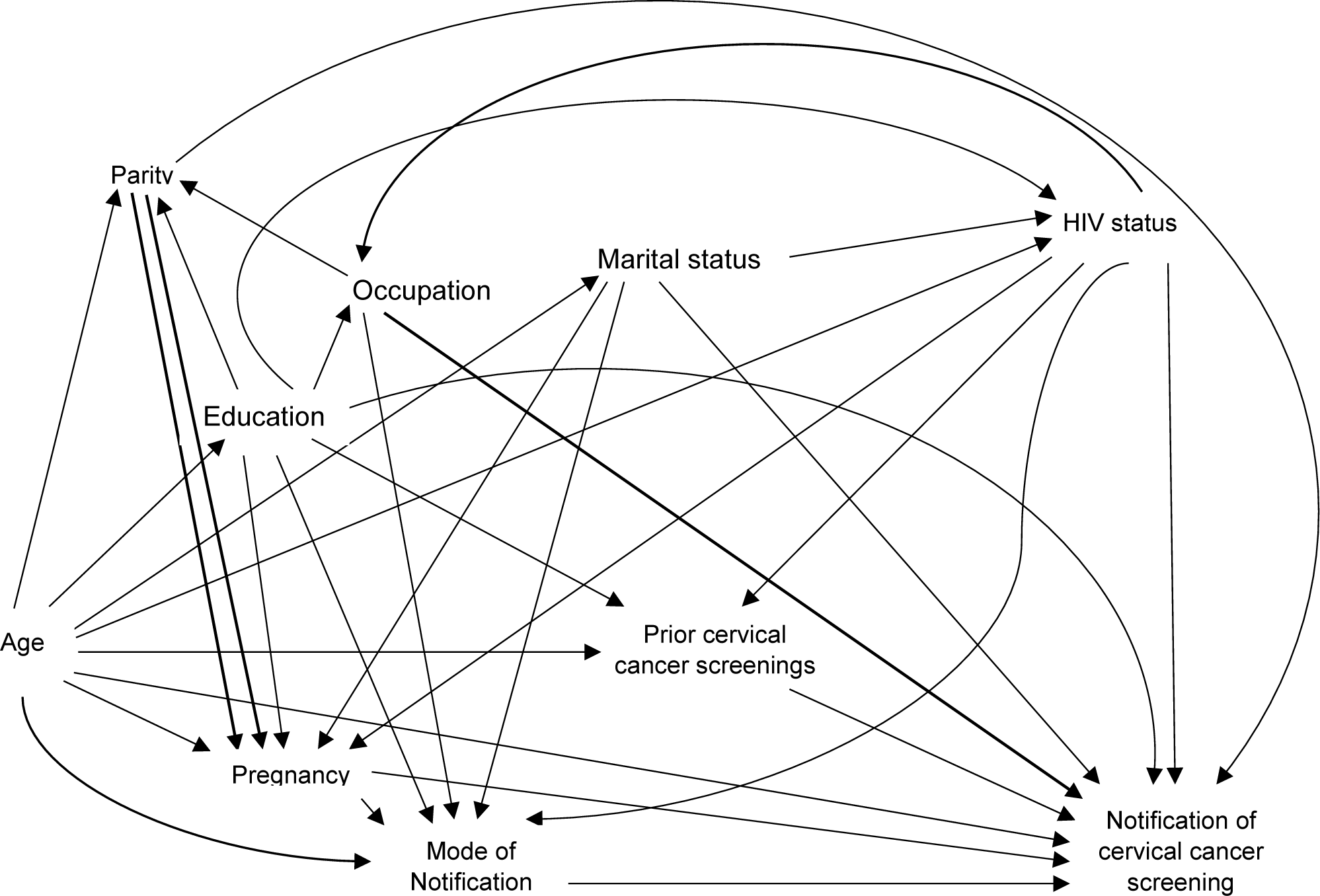
Representative directed acyclic graph (DAG) depicting our hypothesized system under investigation. We sought to explore the independent causal contribution of age, marital status, education, occupation (proxy for socioeconomic status), parity, pregnancy, mode of notification, prior cervical cancer screening and HIV infection status on the ability to notify women of their results from cervical cancer screening performed in a community-based health fair in rural Uganda. Results notification was one aspect of the overall public health approach to cervical cancer screening that also featured community-based screening and community-based treatment by mobile treatment units. The DAG was used to guide which variables to control for when assessing the independent contribution of the various constructs on successful notification. This particular DAG shown focuses on the role of the participant’s preferred mode of notification (via text message, personal phone call, at-home visit by a Village Health Team member, or participant’s visit to a nearby healthcare facility) as the primary exposure variable of interest in determining whether the screening test result was communicated back to the participant.

## Results

### Community-based screening fairs

Between March and November 2016, 2,142 women attended one of 24 health fairs in one of 15 communities in two rural districts in western Uganda and expressed interest, by virtue of registering, in being screened for cervical cancer. The average attendance on each day of fair was about 120 women (Figure 4). Two hundred and forty women were ineligible due to age (96%), symptoms (confirmed by pelvic exam) suggestive of cervical cancer (2.5%), or prior hysterectomy (1.3%). Of the 1,902 women who were eligible and consented for cervical cancer screening, 1,892 (99%) provided a self-collected vaginal specimen (Figure 4). The median age of the participants was 34 years (interquartile range [IQR] 28-40). Majority (79%) had only primary level education and described their job as non-professional work (which included farming, fishing, housekeeping and personal business). Of the women who self-reported HIV status, the prevalence of self-reporting HIV was 10%. Majority of the women had never screened for cervical cancer (94%) including those who were HIV positive. Most women (95%) responded that the venue for cervical cancer screening was easily accessible within 2 km of their homes and most (89%) were able to walk to the venue.

**Figure 4.**
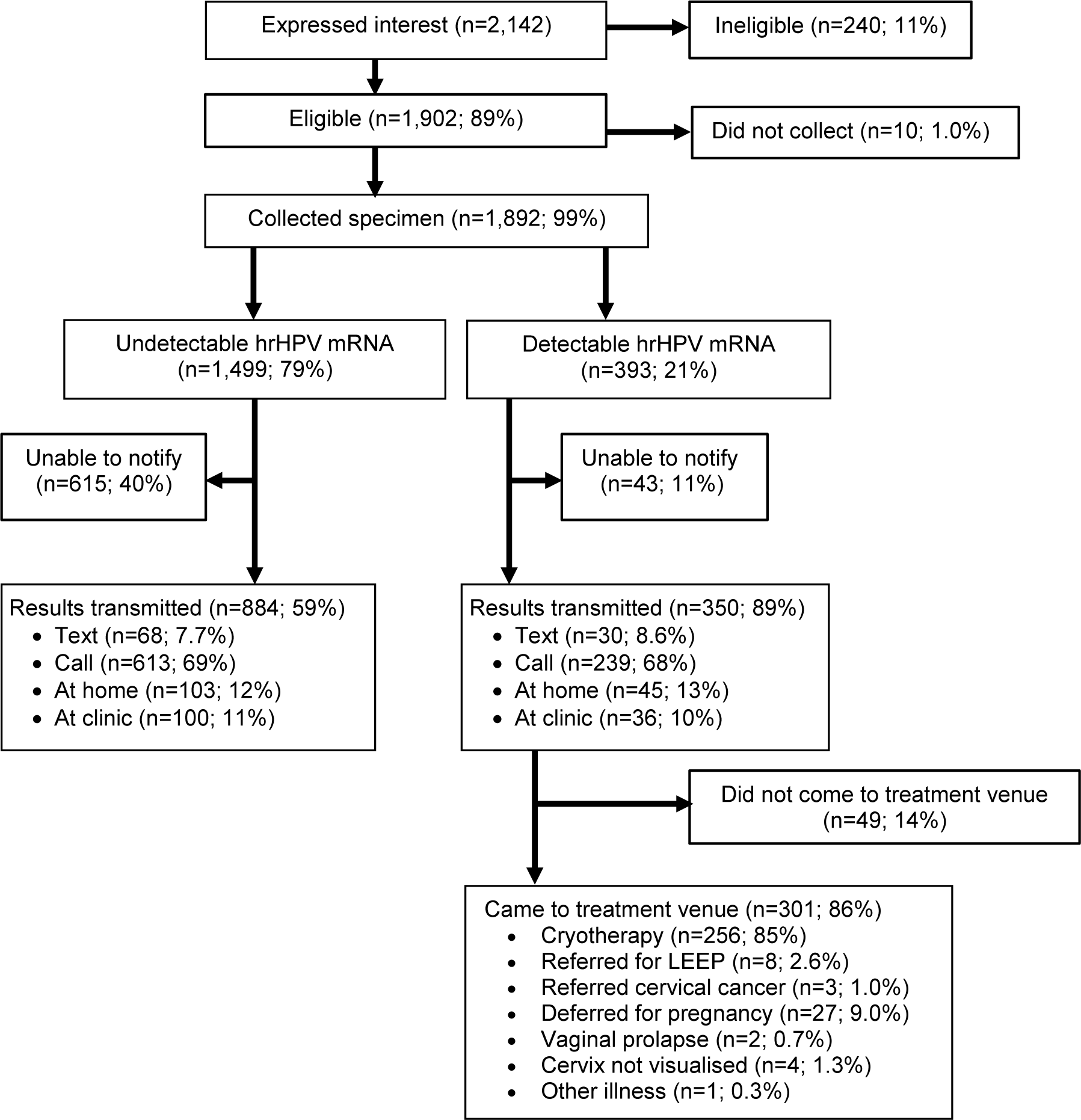
Participation among community women in rural Uganda in various aspects of a public health approach to cervical cancer screening. The approach featured community-based screening, results notification, and community-based treatment by mobile treatment units. LEEP refers to loop electrosurgical excision procedure.

### HPV testing

The women reported to have had a positive experience with self-collection as the majority of them agreed that they would test again and recommend the HPV testing to a friend (Table 1). In addition, most women agreed that the self-sampling instructions were clear and nearly all felt that they had adequate privacy.

**Table 1.**
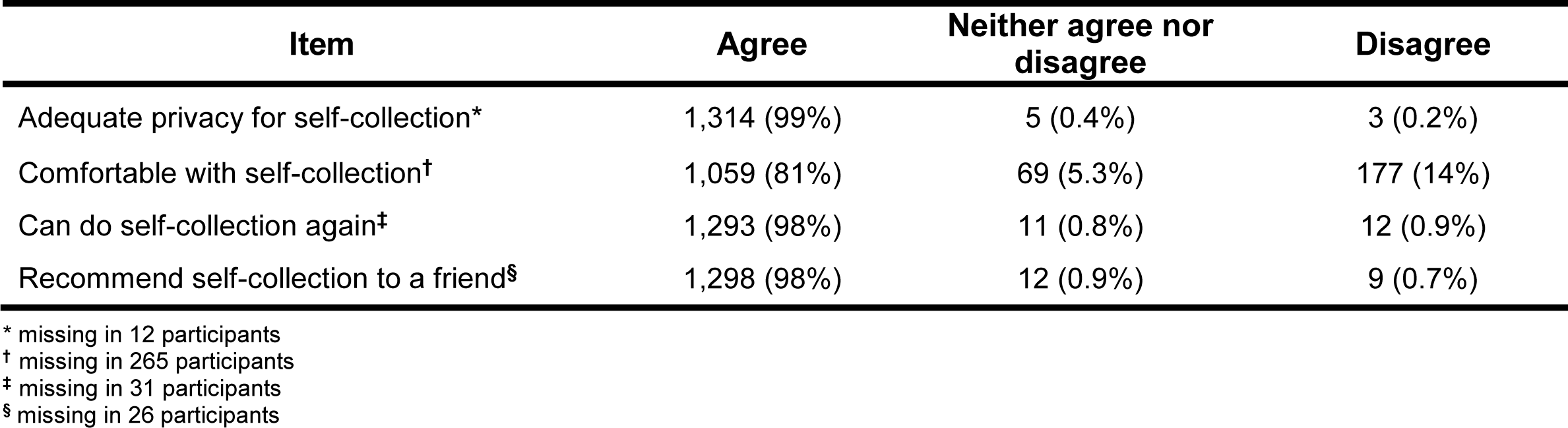
Opinions among women in rural Uganda regarding self-collection of vaginal specimens at a community-based health fair.

### Results notification

Most of the participants (1,877; 99%) indicated a preference for the mode of notification of their HPV testing results. The majority of the participants (77%) preferred to be contacted by phone (70% call; 7.0% text); the remainder preferred coming to a clinic (12%) or a home visit by a VHT (11%). Majority, (66%; 95% CI: 63%-68%) were successfully notified of their results (Figure 5). Of the 393 women who were positive for hrHPV, 350 (85%) were notified, while 884 (59%) of the 1,488 hrHPV negative women were notified. Overall about 734 (60%) of the notified participants were able to be reached on the first attempt at notification: (phone call 425 (58%), SMS 23 (3.1%), home visit 124 (17%), return to clinic 157 (21%). In evaluating independent determinants for successful notification of results, we constructed a directed acyclic graph to depict the system and inform what factors to adjust for (Figure 3). We first evaluated successful notification of results and we found, in unadjusted analyses, that level of education (at least primary and secondary level), notification mode by phone SMS were associated with successful notification. (Table 2). Secondary level education had the largest magnitude of association (unadjusted risk ratio 1.23 95% CI 1.09 to 1.39; P value 0.001. After relevant multivariable adjustment, we found that some education (primary and secondary education), notification mode by SMS remained significantly associated with successful notification while women with non-professional employment were 0.85 times less likely to be notified. Secondary level education still had the largest level of association; (Adjusted risk ratios 1.26 95% CI 1.09 to 1.46 P value 0.002) (Table 2)

**Table 2.** Evaluation of various participant sociodemographic and clinical characteristics for their association with successful notification of results from an HPV test performed at a community-based health fair for cervical cancer screening in rural Uganda.

| Characteristic | No. evaluated | No. (%) successfully notified of results | Unadjusted |  | Adjusted |  |
| --- | --- | --- | --- | --- | --- | --- |
|  |  |  | Risk ratio* (95%CI) | P value | Risk ratio (95% CI) | P value |
| **Age, in years** |  |  |  |  |  |  |
| ≥ 40 | 509 | 337 (66%) | Ref. |  | Ref. |  |
| 30-39 | 760 | 506 (67%) | 1.01 (0.92-1.09) | 0.89 | 0.99 (0.89-1.09)<sup>†</sup> | 0.80 |
| 20-29 | 608 | 391 (64%) | 0.97 (0.89-1.05) | 0.51 | 0.97 (0.86-1.10)<sup>†</sup> | 0.53 |
| **Marital status** |  |  |  |  |  |  |
| Never married | 51 | 35 (69%) | Ref. |  | Ref. |  |
| Married | 1,564 | 1,025 (66%) | 0.95 (0.79-1.15) | 0.63 | 0.95 (0.77-1.18)<sup>‡</sup> | 0.66 |
| Separated/widowed/divorced | 262 | 173 (66%) | 0.96 (0.78-1.18) | 0.71 | 0.95 (0.75-1.19)<sup>‡</sup> | 0.64 |
| **Education** |  |  |  |  |  |  |
| None | 305 | 172 (56%) | Ref. |  | Ref. |  |
| At least some primary | 1,036 | 671 (65%) | 1.15 (1.03-1.28) | 0.012 | 1.19 (1.05-1.35)<sup>§</sup> | 0.006 |
| At least some secondary | 342 | 237 (69%) | 1.23 (1.09-1.39) | 0.001 | 1.25 (1.08-1.45)<sup>§</sup> | 0.004 |
| At least some tertiary | 18 | 12 (67%) | 1.18 (0.84-1.66) | 0.34 | 1.11 (0.76-1.62)<sup>§</sup> | 0.56 |
| **Occupation** |  |  |  |  |  |  |
| Unemployed | 1,499 | 988 (66%) | Ref. |  | Ref. |  |
| Employed, non-professional | 187 | 110 (59%) | 0.89 (0.79-1.01) | 0.08 | 0.86 (0.74-0.99)<sup>¶</sup> | 0.03 |
| Employed, professional | 191 | 135 (71%) | 1.07 (0.97-1.18) | 0.16 | 1.02 (0.90-1.16)<sup>¶</sup> | 0.75 |
| **Mode of notification** |  |  |  |  |  |  |
| Phone call | 1,314 | 852 (65%) | Ref. |  | Ref. |  |
| Home visit | 217 | 148 (68%) | 1.00 (0.90-1.11) | 0.96 | 1.01 (0.89-1.16)<sup>**</sup> | 0.85 |
| Return to nearby clinic | 225 | 136 (60%) | 0.98 (0.85-1.12) | 0.76 | 0.98 (0.86-1.11)<sup>**</sup> | 0.73 |
| SMS<sup>††</sup> | 121 | 98 (81 %) | 1.20 (1.04-1.37) | 0.10 | 1.24 (1.12-1.38)<sup>**</sup> | 0.000 |
| **Pregnant** |  |  |  |  |  |  |
| Not pregnant | 1,687 | 1,098 (65%) | Ref. |  | Ref. |  |
| Pregnant | 190 | 135 (71%) | 1.09 (0.99-1.20) | 0.07 | 1.06 (0.95-1.19)<sup>‡‡</sup> | 0.29 |
| **Parity** |  |  |  |  |  |  |
| 0-3 | 705 | 457 (65%) | Ref. |  | Ref. |  |
| 4-6 | 746 | 485 (65%) | 1.00 (0.93-1.08) | 0.94 | 1.04 (0.94-1.14)<sup>§§</sup> | 0.45 |
| >6 | 426 | 291 (68%) | 1.05 (0.97-1.15) | 0.22 | 1.05 (0.93-1.20)<sup>§§</sup> | 0.42 |
| **Prior cervical cancer screening** |  |  |  |  |  |  |
| Never screened | 1,791 | 1,176 (66%) | Ref. |  | Ref. |  |
| Screened | 86 | 57 (66%) | 1.01 (0.86-1.18) | 0.91 | 0.91 (0.75-1.12)<sup>¶¶</sup> | 0.38 |
| **HIV infection, via self-report** |  |  |  |  |  |  |
| HIV-uninfected | 1,498 | 986 (66%) | Ref. |  | Ref. |  |
| HIV-infected | 158 | 99 (63%) | 0.95 (0.84-1.08) | 0.44 | 1.00 (0.87-1.14)<sup>***</sup> | 0.99 |

\* Risk ratio depicts the probability of successful notification in the index group divided by the probability of successful notification in the reference group.
<sup>†</sup> Adjusted for education, HIV infection status, marital status, notification mode, occupation, parity, pregnancy, and prior cervical cancer screening. Age and notification mode missing in 15 participants.
<sup>‡</sup> Adjusted for age, education, HIV infection status, notification mode, occupation, parity, and pregnancy. Marital status and notification mode missing in 15 participants.
<sup>§</sup> Adjusted for age, HIV infection status, marital status, notification mode, occupation, parity, pregnancy, prior cervical cancer screening. Education and notification mode missing in 191 participants.
<sup>¶</sup> Adjusted for age, education, HIV infection status, marital status, notification mode, parity, pregnancy. Occupation and notification mode missing in 15 participants.
<sup>\*\*</sup> Adjusted for age, education, HIV infection status, marital status, occupation, pregnancy. Notification mode missing in 15 participants
<sup>††</sup> Denotes short messaging service, commonly known as text messaging.
<sup>‡‡</sup> Adjusted for age, education, HIV infection status, marital status, notification mode, occupation, parity. Pregnancy and notification mode missing in 15 participants.
<sup>§§</sup> Adjusted for age, education, HIV infection status, marital status, occupation, pregnancy. Parity and notification mode missing in 15 participants.
<sup>¶¶</sup> Adjusted for age, education, HIV infection status. Prior cervical cancer screening and notification mode missing in 15 participants.
<sup>\*\*\*</sup> Adjusted for age, education, marital status, notification mode, occupation, parity, pregnancy, prior cervical cancer screening. HIV infection, self-reported and notification mode missing in 236 participants.

### Mobile treatment

Of the 350 HPV-positive women who were notified and asked to return for treatment, 301 (86%; 95% CI 82%-89%) returned for the mobile treatment visit (Figure 4). Majority of the women ((85%) were eligible for immediate biopsy and cryotherapy while others were deferred for various reasons that included pregnancy (9.0%), vaginal prolapse (0.7%), inability to safely administer cryotherapy due to cervical position (1.3%), referral to Mulago Hospital for LEEP or suspicion for invasive cancer (3.6%) or concurrent systemic illness (0.3%). Of the 8 women that were referred to Mulago for LEEP, 5 of them complied and received treatment while 2 of the 3 patients with invasive cancer were able to comply and receive treatment. All the three women who were found to have cervical cancer were contacted and referred for treatment.

## Discussion

In a population which had a very low prevalence of prior screening, we implemented a community-based approach featuring VHT, self collected HPV testing, and mobile treatment. One of the first demonstrations of package including VHT-mobilized, self collection, and mobile treatment – all performed very close to where women live in their communities. We demonstrated great interest in screening, acceptability of self-collection, ability to notify women of their results and good compliance with treatment amongst those who were HPV-infected. follow-up. Finally, we found that some education and notification mode by SMS remained significantly associated with successful notification of results while women with non-professional employment were less likely to be notified.

Our program featured community-based screening, VHTs, and self-administered specimen collection. Although WHO recommends protocols that radically simplify the procedures and technologies for screening and treatment, there are additional inherent barriers within resource constrained settings that must be addressed for effective implementation [22] – namely acceptability (self-collection), location and logistics at the screening facilitites as well as education among others. Even amongst women who regularly go to health care facilities, it is dificult for all facilities to maintain screening. For instance, despite the WHO recommendation to integrate cervical cancer screening into existing HIV care services [23], most of our HIV positive patients (92.5%) had never screened even when majority (98%) were in care. We choose to ground our screening in the community because of the axiomatic convenience it provides to women, a belief that has been borne out in several studies comparing community-based screening to health facilities screening in East African countriesand outside East African countries [24-26]. In our approach, we used the VHTM’s within their communities similar to other studies who used the community health work (CHW) force within their communities [24, 26]. Our results show that almost all the women who attended the health fair were able to provide a sample. Indeed other evidence shows that self-collection of vaginal specimens is very acceptable [27], and increases sreening uptake [7, 10] which ultimately increases detection of disease, offsetting the slight decrement in sensitivity [28]. Further still, services may not be offered at that particular time or day when the woman is able to come to the clinic or long waiting time may all increase cost to individuals. Additionally, clinics where screening is offered are often far from a woman’s home and this may limit access due to transportation challenges.

Majority of the women preferred to receive results via the phone when when we asked them to choose among a variety options to learn of their results. The finding of most participants preferring to receive result via the phone is similar to that of a Kenyan study where 71% of the participants preferred receiving their results by phone communication (call or text) [11]. Indeed evidence has proved that mobile phones have become the most accessible form of communication providing information that results in improved health outcomes and/or changed health behaviors [29]. Overall, most of our participants were able to be notified of their results and mainly those who tested hrHPV positive received their results (89%). By design, we tried harder to reach the HPV-infecteds and we were able to reach more of them in actual practice and this is proof of concept that a large fraction can be reached. However, todate, there is still scanty data and the only available study in Uganda that utilized community-based self-collection for HPV testing reported reaching just 47% of their HPV positive by phone [14]. This study was done in an impoverished slumy urban community and this may have had inherent factors that could have contributed to poor access via the phone.

To-date, there is no prior report of mobile treatment offered in a campaign style for cervical cancer. In our study, most of the HPV positive women were able to turn up for their treatment appointments (86%) within the community. The women were facilitated with some modest transport reimbursement to access treatment in addition to the study staff and the VHTs reaching out to them. Our study registered a slightly higher turn up compared to a similar study in Uganda [10] which offered transport reimbursement but, in contrast to our study, held their treatment at larger facilities that were on average likely farther away from participants. Additionally, they did not intensify the tracking down of women to the level of their homes using the VHT’s. Other studies where women were referred to hospitals with no facilitation for transport have even worse return; 39% [26] within East Africa and outside the East African region (59%) [30] and (54%) [31]. Therefore, our turn up could be attributed to the convenient mobile treatment that we set up at low level health facilities within the communities as well as some transprot facilitation. It is of great importance that screening programs minimise LTFU and in this has been the major impetus of any screen and treat approach. Our approach is novel because it brings treatment close to the patient, a scenario that could never be feasible or cost-efficient through service delivery in all local health facilities. For instance, our approach recognizes that it will never be cost – efficient to staff a large number of clinics in a country like Uganda such that most women have such a clinic near them. Instead, our approach creates a very small team of expert personnel who will move across a region or country. Because this medical procedure is never urgent, all clinics do not have to have it. It is fine if the service is only available via a roving team and a few well placed national medical centers. Furthermore, having dedicated treatment days for a large number of known HPV positives will optimize cost-efficiency compared to a model of screen and treat – as it is bound to be erratic based on the prevailing clinic needs.

There are tradeoffs in the public health approach. Indeed, we concede that our approach prioritizes greater community coverage over optimal care for each individual. Specifically; (i) women are told about the need for screening through mass communication as opposed to an individual-level letter or phone call (as would happen in a resource-rich setting) (ii) self-collection of a vaginal specimen is less sensitive than what could be accomplished with clinician-collected cervical specimen (iii) we don’t spend a lot of resources on tracking down those with positive HPV results but whom cannot be notified or those who were notified but do not come back for treatment (iv) we treat all HPV positives, which is certainly over treating many, as opposed to using colpo and biopsy to guide who needs to be treated (v) without a biopsy, we may be missing some cancer and hence undertreating some women by just giving cryotherapy. However, conceding perfection at the individual level is not novel and, in fact, underlies the entire screen and treat philosophy. Therefore, we believe this approach is justified because it allows for a limited amount of resources to reach a larger number of women. Further, what is novel about our approach is that places all of the interventions in one package that is more convenient for women. Generally, this is similar to the WHO public health approach to HIV treatment and care [15] where anti-retroviral therapy was rolled out without plasma HIV RNA testing, resistance testing, or attention to bringing back patients who are lost to follow-up. Some enhancements like same day point of care (POC) HPV testing and treatment may further improve this public health approach to cervical cancer screening. Finally, we are not able to tell exactly how many more women are reached and this awaits a formal decision analysis.

Our study had several limitations. There is no way of knowing penetrance (how many heard the message) and success of our mobilization (percentage who came among those who heard) and why those who did not come did so. Because we don’t know who did not come, it is unknown which women came for screening and if they are a representative sample of the target population. Thus, it is unclear how representative our inferences are for the whole community (e.g., by just questionning those who came we don’t know the percentage who found it convenient to come or the percentage who found self collection acceptable). There was a delay between testing and results notification which may have decreased response percentage. Although the initial mobilization did not mention research, once women arrived at the health fairs they became aware that our demonstration project was a research study. This may have become known to other women which then influenced their choice to come on later days. Participation may be higher when this approach is implemented devoid of research affiliation. Of those who came to the fairs, because our activities were couched in the form of a research study, this may have influenced their answers to questions such as acceptablity of self collection and everything thereafter. In other words, we don’t know how all of this will perform in real world conditions without any mention of research.

In conclusion, the public health problem of cervical cancer requires public health solutions. Our present study has showed that a village health team led community based campaign for cervical cancer screening had a lot of interest in community, acceptablilty of self collection and good retention through to treatment. Our approach can be implemented now: This public health approach to cervical cancer screening should be considered by policy makers and researchers for wider implementation and evaluation to ultimately reduce the burden of cervical cancer in the LMICs. It also merits further optimization by (next steps): understanding and optimizing penetrance and mobilization; overcoming whatever barriers might exist for some women, and optimizing cost-efficiency at every step.

## Data Availability

Data available on request from the authors

## Acknowledgement

We gratefully acknowledge Jane Francis Nalubega, Proscovia Kyomuhangi, Mary Mubeezi and Patricia Nabayego for their implementation of the field work, and Frank Mubiru for statistical support. We also thank the VHTs and Jive Media Africa (Robert Inglis) for the mobilization campaigns.

## Abbreviations

CI: confidence interval
DAG: directed acyclic graph
HIV: human immunodeficiency virus
HPV: human papillomavirus
hrHPV: high-risk human papillomavirus
IQR: interquartile range
mRNA: messenger ribonucleic acid
VHTs: Village Health Team members
WHO: World Health Organisation

